# Evaluation of SURUS: a Named Entity Recognition System to Extract Knowledge from Interventional Study Records

**DOI:** 10.1101/2024.05.31.24308278

**Authors:** Casper Peeters, Koen Vijverberg, Marianne Pouwer, Bart Westerman, Maikel Boot, Suzan Verberne

## Abstract

**BACKGROUND:** Medical decision-making is commonly guided by systematic analysis of peer-reviewed scientific literature, published as systematic literature reviews (SLRs). These analyses are cumbersome to conduct as they require large amounts of time and subject matter expertise to be available. Automated extraction of key datapoints from clinical publications could speed up the process of systematic literature review assembly. To this end, we built, trained and validated SURUS, a named entity recognition (NER) system comprised of a Bidirectional Encoder Representations from Transformers (BERT) model trained on a highly granular dataset. The aim of this study was to assess the quality of classification of critical elements in clinical study abstracts by SURUS, in particular the patient, intervention, comparator and outcome (PICO) elements and elements of study design.

**DATASET & METHODS:** The PubMedBERT-based model was trained and evaluated using a dataset of 400 interventional study abstracts, manually annotated by experts using 25 labels with a total of 39,531 annotations according to a strict annotation guideline, with Cohen’s κ inter-annotator agreement of 0.81. We evaluated in-domain quality, and assessed out-of-domain quality of the system by testing it on out-of-domain abstracts of other disease areas and observational study types. Finally, we tested the utility of SURUS by comparing its predictions to expert-assigned PICO and study design (PICOS) classifications.

**RESULTS:** The SURUS NER system achieved an overall F1 score of 0.95, with minor deviation between labels. In addition, SURUS achieved a NER F1 of 0.90 for out-of-domain therapeutic area abstracts and 0.84 for observational study abstracts. Finally, SURUS showed considerable utility when compared to expert-assigned PICOS classifications of interventional studies, with an F1 of 0.89 and a recall of 0.96.

**CONCLUSION:** To our knowledge, with an F1 score of 0.95, SURUS ranks among the best-performing models available to date for conducting exhaustive systematic literature analyses. A strict guideline and high inter-annotation agreement resulted in high-quality in-domain medical entity of a finetuned BERT-based model, which was largely preserved during extensive out-of-domain evaluation, indicating its utility across other indication areas and study types. Current approaches in the field lack the granularity in training data and versatility demonstrated by the SURUS approach, thereby making the latter the preferred choice for automated extraction and classification tasks in the clinical literature domain. We think that this approach sets a new standard in medical literature analysis and paves the way for creating highly granular datasets of labelled entities that can be used for downstream analysis outside of traditional SLRs.

## Introduction

Interventional trials are an important source of scientific data for medical decision-making. Unstructured data from trials are carefully evaluated in systematic literature reviews (SLRs), which are typically accompanied by meta-analyses. These efforts result in essential medical documents that help drive decision making in the medical field. The key purpose of SLRs is to provide scientific validity through the inclusion of the complete body of evidence to answer a specific research question through an evidence-based approach. As such, the generation of SLRs is an intricate process, during which a broad selection of literature in the field of study is manually screened for eligibility and evaluated for the quality of evidence. It is paramount that an SLR represents an exhaustive evaluation of an area within a scientific field, and it is of high importance that the task of eligibility assessment is scrutinized and performed with complete recall to avoid incorrect exclusion of evidence. This assessment is particularly important in the context of the growing body of scientific publications available; from 1960 onwards, the number of PubMed records has grown exponentially [1], [2]. Nowadays, the initial screening process for SLRs in active medical fields often includes more than 3000 scientific abstracts. This means that assuming one would be able to process abstracts at a pace of 1 per minute, this task alone would take a human at least 50 hours of reading time. Given that the Cochrane Institute’s guidelines for an SLR involve independent screening by two trained experts, a modern, manual SLR process places a disproportional workload on expensive medical experts, often resulting in months of full-time work and costs easily exceeding $100,000 per SLR [3], [4].

Clinical questions for evidence-based practice are typically structured according to elements of an established framework called PICO. For a clinical SLR project, information on the <u>P</u>atient, Intervention, <u>C</u>omparator, and <u>O</u>utcome (PICO) are defined to determine the scope of the work and the trials eligible for the project [5]. For example, the PICO ^1^ framework of a trial could comprise “acute coronary syndrome” (<u>P</u>atient), “rivaroxaban” (Intervention), “placebo” (<u>C</u>omparison) and “systolic blood pressure” (<u>O</u>utcome). Along with elements listed in the PICO framework, study design characteristics (“randomized”) provide additional valuable insights for the selection of eligible studies [6], [7]. Hereafter, the combination of PICO and study design characteristics will be referred to as the PICOS framework or PICOS in short.

One of the challenges in the identification of elements of PICOS is their dependence on textual context. For example, “stroke” may refer to a criterium of study participants for their inclusion into the study (i.e. part of “<u>P</u>atient”) or to an endpoint that is measured during the study (i.e. part of “<u>O</u>utcome”). It may also refer to related research, in which case its identification is of no use to the reviewer.

Over the past decades, the increasing popularity of machine learning (ML) models has given rise to the development of methods to speed up the SLR screening process. Some ML approaches rank scientific publications according to their eligibility to a research question, thus providing the reviewer with the option of a priority cut-off for screening [8], [9], [10]. Alternatively, ML methods can provide the reviewer with information on scientific publications, which can be used to include or exclude studies in further analyses. In particular, ML-based natural language processing (NLP) methods may extract elements of PICOS from unstructured medical text or predict the eligibility of a study based on a set of eligible studies initially selected by the reviewer. Ultimately, accurate and complete extraction of study characteristics by ML models could enable reviewers to base their eligibility decisions on the model outputs during the screening process of an SLR. A subset of biomedical NLP methods currently focuses on named entity recognition (NER) classification techniques. Using NER, unstructured text is processed and words, expressions or sentences are labeled with pre-defined classes (e.g. diseases, drugs, etc.) [2].

Several approaches have been proposed for the extraction of elements of PICOS from clinical publication texts [11]. Despite their apparent advantages, these NLP tools currently have a few limitations: (1) valuable study design features are often not extracted (e.g. study duration and study size); (2) PICO-focused ML-solutions typically focus on prediction of relatively large text sequences, resulting in coarsegrained extraction of limited use to the reviewer; (3) the quality of current NER systems are insufficient to approximate expert reviewer eligibility assessment performance [12]; and (4) there are only few datasets available which are designed specifically for PICO extraction [13], [14], [15], [16], but they are limited in terms of size and granularity, and models trained on these datasets lack performance required.

In this paper, we provide an elaborate evaluation of SURUS, a BERT-based classification model finetuned on a highly granular, manually annotated dataset of medical annotations. SURUS was designed for the extraction of PICOS elements from clinical texts. SURUS (which is not an acronym) was trained to classify 25 different annotation labels in the abstracts of interventional studies. In addition to this, the SURUS NER method design is intended to facilitate the extraction of the results of clinical endpoints using relation extraction. Currently, SURUS is being integrated in software for systematic literature selection and analysis by medical professionals and scientists. The software allows for identification of relevant literature through the recognition of medical named entities and abstract sections.

The primary aims of this research were to validate the annotation method used to create the fine-grained dataset underlying SURUS and to evaluate the quality, consistency and utility of the SURUS system. Our contributions are two-fold: (1) we show that, using a detailed and highly granular annotation method, the BERT model can be fine-tuned to recognize a large diversity of contextually divergent medical entities with high accuracy; (2) the annotation method proposed can be used for classification of key, medically relevant concepts in clinical texts with high reliability, and may be extrapolated for use in other (medical) fields. To the best of our knowledge, SURUS is the first deep learning-based system capable of extracting a wide variety of clinically relevant entities from a text with an accuracy high enough for use in clinical practice.

### Related work

Previously, data classification and categorization of scientific study records were experimented with using Support Vector Machines (SVMs), Conditional Random Fields (CRFs), Long Short-Term Memory (LSTM) or, more recently, Bidirectional Encoder Representations from Transformers (BERT) models. Examples of tools employing one or more of these techniques were recently reviewed and evaluated in a systematic review [11].

Whilst all of the models listed above have their advantages and drawbacks, the consensus is that transformer-based methods such as BERT combine high potential with relatively small (annotation) effort compared to alternatives [11]. BERT is a transformer encoder model, pretrained on a vast dataset of books and Wikipedia [17]. BERT models have shown superiority compared to BiLSTM models on several tasks including NER [18], [19]. Later on, BERT was expanded upon by adding biomedical scientific texts to its pretraining, including the specialized BERT-derivatives BioBERT [20], SciBERT [21] and PubMedBERT [22].

The most recent innovation in the field of NLP are generative large language models (LLMs). LLMs, such as GPT-4 and ALPACA, excel in summarization, contextualization and extrapolation of information from a wide range of scientific fields. In the medical field, contributions include summarization of medical texts, chat-bot mediated diagnosis and medical education [23], [24]. However, the generative nature of these models make them prone to hallucination and classification inaccuracy, which is undesirable in a task demanding extensive classification recall [23], [25]. For this reason, we decided to use a variant of BERT as the classification model of choice for validation of the quality of the dataset presented here.

BERT-based models can be fine-tuned to perform well in specialized supervised learning tasks. Manual, task-specific labeling for fine-tuning a model is work-intensive and requires expert knowledge of the task and domain. In addition, currently available datasets for NER are often of limited quality and consistency [26].

To our knowledge, there are currently 3 datasets publicly available for recognition of PICO specifically. First, Kim *et al*. created the NICTA-PIBOSO dataset^2^, which consists of 1000 abstracts with manually labeled sentence annotations amongst 5 label classes [27]. Second, Jin *et al*. presented the PubMed-PICO dataset^3^, consisting of almost 25,000 abstracts of which relevant sentences were automatically assigned to 1 of 7 labels using a rule-based algorithm [28]. Third, Nye *et al*. [29] reported the EBM-NLP corpus^4^, which consists of 5190 abstracts of scientific publications, 190 of which are annotated by experts and 5000 by laymen, using Population, Intervention and Outcome labels. The EBM-NLP corpus was used to train PICO-extracting systems on a sentence [30], [31] and span level [32], [33].

For SURUS to be able to accurately and concisely predict elements of PICOS, we created a dataset that offers the following advantages: (1) the annotation approach is suitable for word-level extraction; (2) we distinguish 25 different labels allowing for fine-grained extraction of PICOS characteristics; (3) the dataset presented here consists exclusively of expert-annotated labels; (4) our dataset is designed in a way that would facilitate extraction of detailed study results through relation extraction in a later phase of SURUS development.

## Materials and Methods

### Dataset

For our dataset, we used a set of scientific articles abstracts, publicly available in the PubMed database^5^. PubMed is the most widely used source of clinical evidence and consists of the Medline and PMC databases. Our dataset consisted of abstracts of interventional study reports. Interventional studies are characterized by investigation of a medical intervention and group distinction is typically based on differences in therapeutic regimen [34]. Though of similar study type, the style of reporting may vary greatly between therapeutic areas and interventional study subtypes. For this reason, abstracts included in the SURUS dataset were of various interventional study subtypes and therapeutic areas. T o ascertain high versatility, 4 of the most important therapeutic areas as reported in WHO ICD-11^6^ were selected to be represented in the dataset: cardiovascular diseases, endocrine diseases, neoplasms and respiratory diseases. In total, 400 article abstracts of interventional studies (100 for each therapeutic area) were randomly selected from the PubMed database for in-domain evaluation of the NER system. In addition, a set of 123 other article abstracts was randomly selected for out-of-domain therapeutic area (90) and study type (33) evaluation. During randomization, the aim was to achieve a fitting representation of the real-world diversity of interventional publication abstracts in our dataset.

#### Expert annotations in the NER dataset

The abstracts of these selected publications were manually annotated. During annotation, entities were labeled and assigned to one of 25 labels, amongst 7 label classes. Label classes that not relevant to extraction of PICOS elements were designed for either extraction of additional valuable information outside of PICOS or extract entities of study results. In addition, an element of PICOS may consist of multiple labels. For example, “Population” may be composed of entities of the “Methodology Inclusion Criteria” but also “Disease Indication”. We chose this structure to clearly define the contextual niche of every label class, and to add to the granularity and the utility of the predictions made. All label classes had distinct contextual dependencies and unique labels. A full overview of annotations in the dataset is visualized in Figure 1, the mapping of labels to elements of PICOS and more detailed descriptions of the label class are available in Appendix table B. Correct labeling of text elements is dependent on the context of the element and annotations made in its vicinity. For example, when mentioned in the methods section, “overall survival” was labeled as an element of the label class ‘Methodology’, whereas it was labeled as an element of the ‘Parameter’ class in the results section. However, when “overall survival” was mentioned in the results section without any association with annotations of the ‘Result’ class (so without associated results), it was not labeled at all. These nuances add to the intricacy of the annotation process.

**Figure 1.**
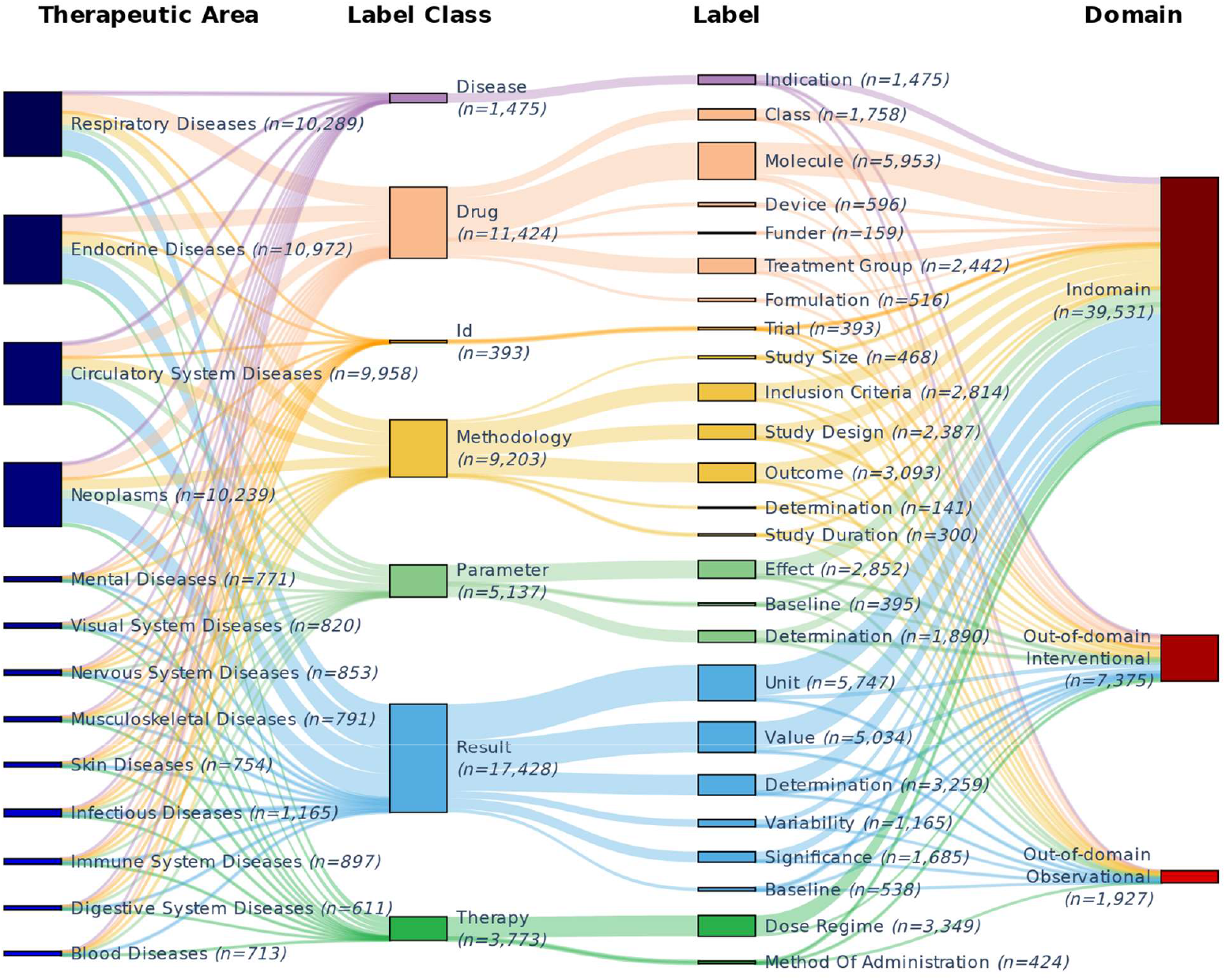
Sankey diagram of all 48,833 expert annotations in the NER dataset. From left to right, the distribution of annotations is illustrated between different categories (nodes) of therapeutic area, annotation label class, annotation label and record domain. The width of the connections between nodes illustrate the number of overlapping annotations between nodes. Total number of annotations in nodes are listed between brackets behind the node name.

In total, the 400 scientific abstracts were labeled with 39,531 annotations, averaging 98.83 (±29.70) annotations per abstract.

#### NER dataset annotation process

Master students with a pharmaceutical or biomedical background were tasked to annotate the scientific abstracts. To warrant the quality and consistency of the annotations made, we made four provisions: (1) a detailed annotation manual was assembled by the first author to guide the annotators; (2) all annotators followed a 2-day course, during which they were instructed about the annotation methodology and process; (3) all annotations made were reviewed by one of two expert annotators; (4) annotation consistency was manually monitored using an extensive set of restrictive rules for annotation span range and context.

The primary aims of the manual created was to facilitate complete extraction of PICOS elements and to promote consistency of annotations made between articles in different fields and of different designs. Due to the high diversity of contextual situations in medical articles, the assembly of the manual was an iterative process featuring regular ‘consensus sessions’, during which the judgment of one expert annotator was decisive. Figure 2 shows a fully annotated example of a study abstract.

**Figure 2.**
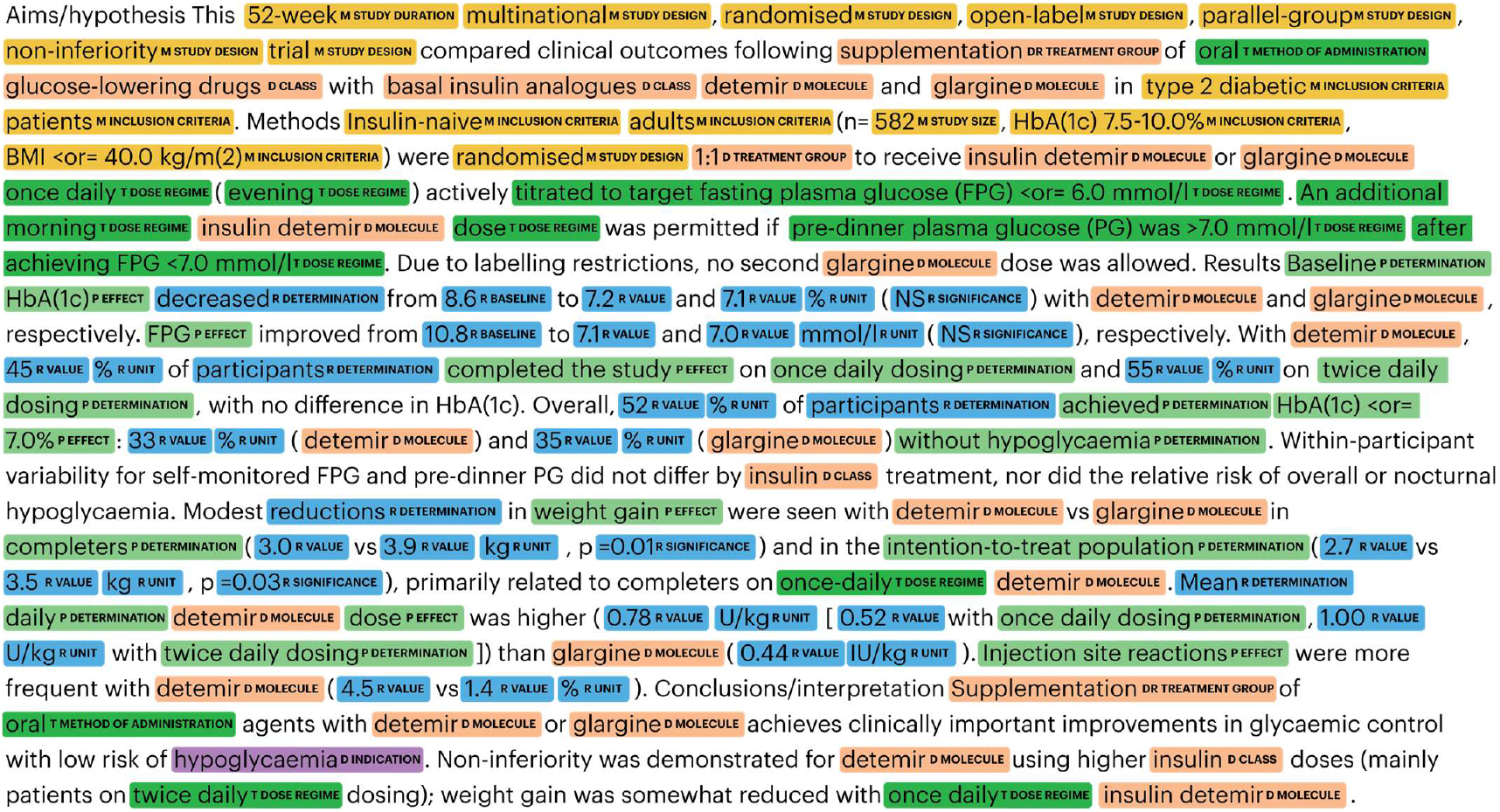
An example of a fully annotated interventional study abstract of the record with Pubmed identifier 18204830. Different types of labels are colored according to their label class. The label class of different annotations is abbreviated in the image. For example, “P EFFECT” represents an ‘Effect’ label of the ‘Parameter’ class.

#### Inter-annotator agreement

To estimate the reliability of the data, we measured inter-annotator agreement (IAA) between four annotators (two expert annotators and two of the student annotators) on a randomly determined subset of the scientific abstracts. For IAA assessment, 35 scientific abstracts (5 for each therapeutic area in the annotated dataset) were randomly selected to be separately annotated by the four annotators. Due to the abundance of unlabeled tokens in the dataset (introducing a positive bias), F1 on a token level was calculated in addition to Cohen’s κ to approximate IAA, as unlabeled tokens may be left out of the calculation with this method [35], [36], [37], [38]. Token-level agreement between annotators had a κ of 0.81 (±0.05 between articles) and a F1 of 0.88 (±0.01).

#### Dataset split into test and train sets

After annotation, all (n=400) abstracts were split randomly into 10 partitions, each consisting of 10 articles from each therapeutic area (Appendix table C), which were used for model quality evaluation through k-fold cross-validation. Randomization of the abstracts was stratified by the presence of headers in the abstract, annotation-to-word ratio and the number of study arms. The Randomice tool was used for unbiased randomized stratification of records amongst the datasets [39].

### Model

#### Input for the NER system

All abstracts of the NER dataset were tokenized using the BERT tokenizer^7^ and subword token embedding tensors were assigned with the BERT base uncased model^8^. It is common for clinical publication abstracts to consist of more than 512 subword tokens. To resolve this issue of exceeding the BERT input limit of 512 subword tokens, we used a sliding window approach. Scientific abstracts longer than 512 subword tokens were divided into *n* batches of 512 sub-word tokens, with a 256 subword stride. The number of batches (*n*) was determined according to:

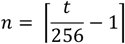

Where *t* was the total number of subword tokens. For example, a scientific abstract of 1200 subword tokens was divided into 4 batches.

#### NER model training

The NER model was trained on all train set articles of the NER dataset. 512 subword tokens at a time were fed to BERT in the sliding-window approach. For training, a learning rate of 5*10^−5^ (momentum 0.99) with Adam optimization was used, training for 8 epochs using a batch size of 1. The system was trained to assign a BILOU tag and one of 25 labels, based on BERT prediction. Compared to more conventional BIO tags, BILOU tags (Beginning, Intermediate, Last, Outside, Unit) allow for more granularity of a dataset by distinguishing between single- and multiple-token chunks [40]. In the sliding window set-up, a BILOU tag and label of a subword could be predicted up to 2 times (the label predicted may differ between predictions, due to the context difference between strides). During post-processing, the average of the probabilities for each label predicted between batches was taken as the final prediction, and the label with the highest probability was assigned to the token. Finally, adjacent tokens with the same annotation label were aggregated into a single annotation according to their BILOU classification pattern.

#### Quality evaluation

Evaluation of the model quality was done by calculation of the precision, recall and F1 (Eq. 1) of the model output compared to annotations in the test set. NER evaluation was done on the entity level with only complete matches as true positives. A complete match was defined by a token start, token end and label match between predicted and true labels. As such, the corresponding label class but different prediction onset or end compared to the annotation was insufficient for a complete match. For example, a span classified by NER as ‘Inclusion Criteria’ and annotated as ‘Outcome’ did not yield a full match, even though both are of the ‘Methodology’ label class. Similarly, comparison of a prediction of “complete remission” with an annotation of “remission”, both in the ‘Effect’ label, yielded a false positive.

**Eq. 1-** Equations describing calculations of precision (left), recall (middle) and F1 measure (right) using true positives (tp), false positives (fp) and false negatives (fn).

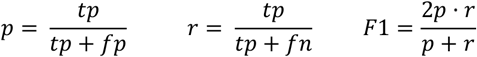

### Experiments

The system quality was evaluated in two settings: in-domain and out-of-domain quality. All in-domain metrics reported were the result of 10-fold cross-validation. Quality assessment was based on the F1 mean and standard deviation over the different labels resulting from the set of measurements. First, we present the experimental setup of in-domain evaluation of the NER and section prediction systems. Subsequently, we describe experiments concerned with consistency and out-of-domain quality. Finally, we describe the protocol of a utility study comparing expert PICOS annotations with the system.

### In-domain quality evaluation

For evaluation of in-domain system quality, the F1 measure of the system was evaluated on a test set of abstracts describing a similar therapeutic area. Evaluation of in-domain quality consisted of four phases: (1) the optimal BERT model for the task was selected through experiments; (2) the quality of the section prediction system was measured; (3) its added value to the F1 of the NER model was evaluated; and (4) using the optimal model, the robustness of the dataset was evaluated.

First, the optimal BERT model to be used during further experimentation was determined. NER quality of four pretrained BERT models (BERT base and domain-specific alternatives BioBERT [20], SciBERT [21] and PubMedBERT [22]) was tested through 10-fold cross-validation, using the train-test splits of all 400 annotated abstracts as specified in section 2.1.4. Based on the resulting F1, the best performing model was selected to be used in the remainder of the experiments. The selected optimal model was the one with the highest mean F1 score between runs.

We assessed the effect of a smaller training set on the in-domain NER prediction quality of the optimal NER system. Prediction quality was compared between systems using 2, 3, 4, 5 and 7 batches as training set (each batch consists of 10% of all dataset articles). This was done using 10-fold cross-validation, where each training fold consisted of block number *k* as the testing set and block numbers *[k+1…k+n+1]* as the training set where *k* was the fold number and *n* was the number of batches included in the training set.

### Out-of-domain quality evaluation

We assessed the quality of the SURUS for abstracts either on another subject or of a different type than the ones included in the annotated training set. For this, we tested the performance on two out-of-domain test sets: one on out-of-domain therapeutic areas and another one on out-of-domain observational study types. For each out-of-domain NER experiment, the SURUS system was tested on abstracts manually annotated by experts as out-domain test sets, according to the annotation rules applied during the annotation of the in-domain dataset. In the out-of-domain therapeutic area test set, we randomly included 10 article abstracts from 9 ICD-11 therapeutic areas not included in the in-domain dataset. In the out-of-domain observational study type dataset, we randomly included 33 abstracts of various observational study types. Amongst the observational study types of the included articles were cohort studies, case-control studies, diagnostic accuracy studies and case studies. Abstracts included in type out-of-domain quality evaluation were of the same therapeutic area as the ones included in the SURUS dataset. A detailed overview of the composition of the out-of-domain NER datasets is provided in Appendix table C.

### Utility of SURUS

To determine the utility of the dataset in the workflow of a systematic literature review specialist, we compared SURUS predictions to expert-determined PICOS characteristics of interventional studies. For this evaluation, we worked with elements of PICOS from Cochrane published in a systematic literature review. 8 study records (2 for each therapeutic area included in the dataset) were randomly picked from 8 Cochrane systematic literature reviews. The Cochrane-assigned elements of PICOS were extracted from the “Characteristics of studies” section. Any element of study design or patient eligibility of the included studies mentioned in the methods section of the Cochrane review was also added to the experiment. Elements of intervention and comparison were merged as these show very limited contextual differences.

To appropriately compare Cochrane classifications to SURUS predictions, two preparatory steps preceded the comparison:

1. All Cochrane-determined elements were manually screened for presence in the study abstract. Any element not present in the abstract was excluded from the experiment. This step was included because Cochrane experts make use of the full record rather than the abstract to determine elements of PICOS.
2. SURUS predictions were mapped manually to Cochrane-assigned elements, as Cochrane-assigned elements may use different wording compared to the abstracts,. The full mapping for the experiment is documented in Appendix table D.

After these steps, the precision, recall and F1 of the SURUS predictions were calculated. For these calculations, the metrics were defined as follows:

- *True positives* were unique predictions correctly mapped towards the correct constituent of PICOS.
- *False positives* were unique predictions that are either not mapped or mapped to the wrong element of PICOS.
- *False negatives* were elements of PICOS to which no prediction of SURUS was mapped or for which elements of SURUS inadequately describe the content.
- *True negatives* were not included in the evaluation as these would bias the quality evaluation.

### Availability

The full code for NER training, the full NER dataset and the detailed annotation guideline for reproduction efforts will be publicly available upon journal publication of the manuscript.

## Results

We report the results of experiments regarding the quality, robustness and out-of-domain viability of SURUS. The experimental results are listed in the following order: results of the in-domain evaluation (1); results of out-of-domain evaluation (2); results of a utility case-study (3). Recall, precision and support for all classes of all evaluations are listed in Appendix table D.

### PubMedBERT performs superior compared to other BERT variants when fine-tuned on SURUS dataset

To determine the optimal BERT model for SURUS, we compared the F1 using BERT, BioBERT and PubMedBERT on the full NER dataset. BioBERT and PubMedBERT showed similar prediction quality overall with an F1 of 0.95, as well as for the predictions of entities from different label classes. The results of the evaluations are listed in Table 1 and more detailed result metrics are listed in Appendix table A. Both models improved NER F1 compared to BERT for all annotation classes and compared to SciBERT for most label classes. The finetuned NER systems showed high prediction accuracy for Drug and Methodology, the label classes most commonly featured in PICOS.

**Table 1.** F1 scores and standard deviations between folds of the 10-fold cross-validation of NER with BERT and 3 science domain-specific derivatives BioBERT, SciBERT and PubMedBERT.

| Label Class | BERT | BioBERT | SciBERT | PubMedBERT |
| --- | --- | --- | --- | --- |
| Disease | 0.92 (±0.03) | 0.95 (±0.02) | 0.94 (±0.01) | 0.95 (±0.02) |
| Drug | 0.93 (±0.01) | 0.95 (±0.02) | 0.95 (±0.01) | 0.95 (±0.01) |
| Identifier | 0.95 (±0.02) | 0.97 (±0.03) | 0.97 (±0.02) | 0.98 (±0.01) |
| Methodology | 0.91 (±0.02) | 0.94 (±0.01) | 0.93 (±0.01) | 0.94 (±0.01) |
| Parameter | 0.81 (±0.04) | 0.87 (±0.02) | 0.86 (±0.03) | 0.87 (±0.03) |
| Result | 0.96 (±0.01) | 0.98 (±0.00) | 0.98 (±0.00) | 0.98 (±0.00) |
| Therapy | 0.90 (±0.03) | 0.93 (±0.03) | 0.93 (±0.03) | 0.94 (±0.02) |
| Weighted Average | 0.92 (±0.01) | 0.95 (±0.01) | 0.94 (±0.01) | 0.95 (±0.01) |

BioBERT and PubMedBERT performed superior compared to BERT and SciBERT. We expected that the performance of a PubMedBERT-finetuned model would extrapolate better for an out-domain task compared to a BioBERT-finetuned model, considering its specialization on Pubmed texts. For this reason, we decided to use PubMedBERT for the remainder of the dataset validation.

### Prediction quality plateaus at training on 70% of dataset items

To assess the rigidity of the annotation method, and the feasibility of further improving F1 by adding more training data, we fine-tuned the SURUS model leaving out varying percentages of the training set. High prediction quality was reached using a small selection of training data (F1 >90% using 20% of the dataset for training, Figure 3). For all categories, F1 mean and variability increased gradually with increasing dataset use, with the highest F1 and lowest variability eventually reached using the full train set (90% of the dataset).

**Figure 3.**
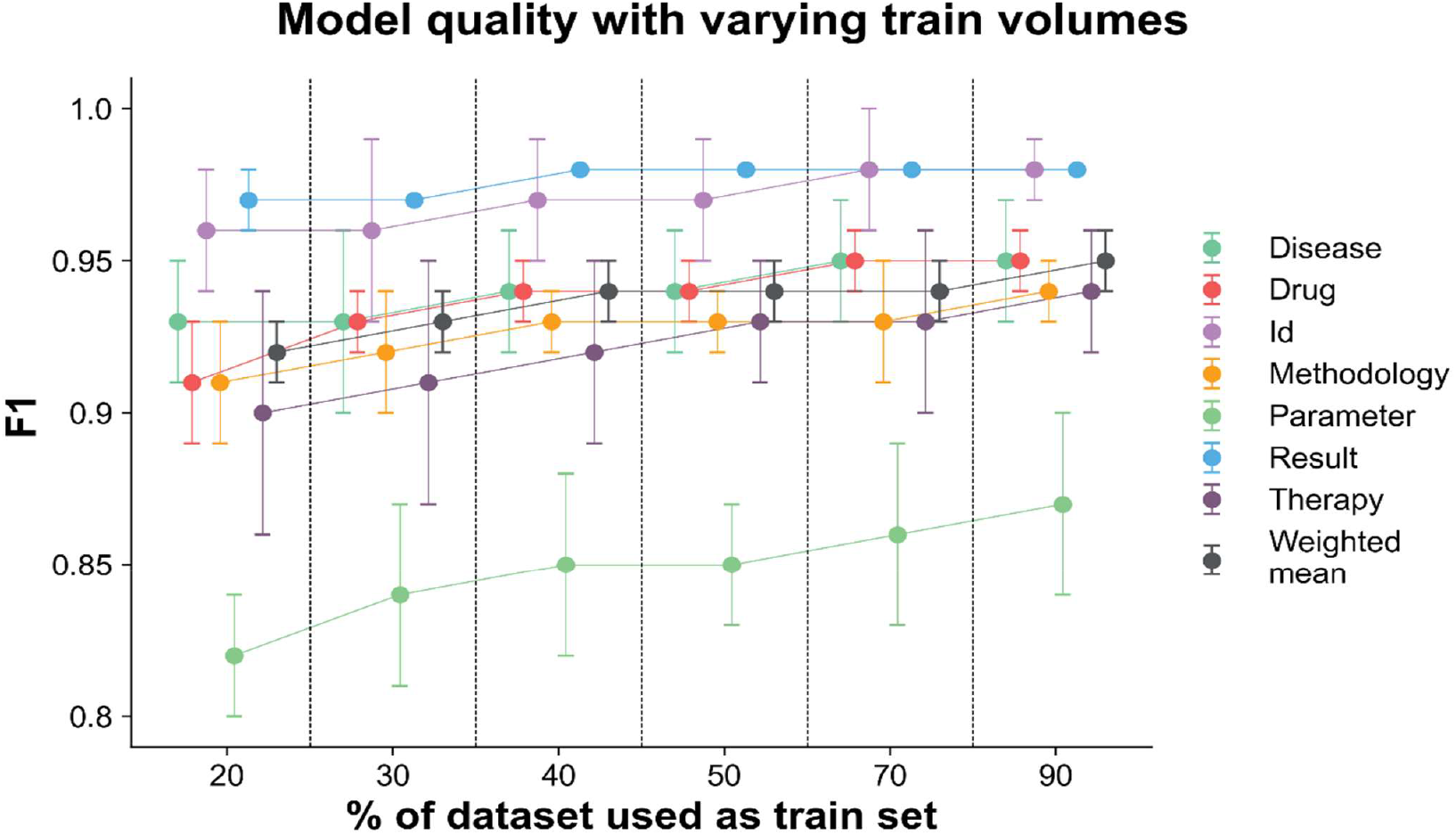
Effect of limiting the volume of train data on the model quality. Weighted mean F1 does not dip below 0.9 even when 80 annotated abstracts are used for finetuning. Mean F1 steadily increases up to 0.95 with full use of train corpus (90% of the dataset). Individual label classes show a similar trend, with a relatively steep increase in context understanding for the Parameter label class, improving up to 0.05 in F1.

### Prediction quality was largely upheld testing out-of-domain abstracts

To evaluate the feasibility of using the system on other types of abstracts than the ones included in the dataset, we assessed the F1 on abstracts of out-of-domain therapeutic areas and observational study type (Table 2). The F1 of the finetuned model on the out-of-domain therapeutic area dataset was 0.90. Similar to the in-domain evaluation, prediction of the Parameter label class appeared to be most inconsistent in the observational dataset relative to the other label classes (the out-of-domain study type), the model scored an overall F1 of 0.84.

**Table 2.** Out-of-domain evaluation metrics of PubMedBERT finetuned on the full SURUS dataset.

| Label class | Interventional |  |  |  | Observational |  |  |  |
| --- | --- | --- | --- | --- | --- | --- | --- | --- |
|  | Precision | Recall | F1 | Support | Precision | Recall | F1 | Support |
| Disease | 0.99 | 0.90 | 0.94 | 664 | 0.95 | 0.87 | 0.91 | 302 |
| Drug | 0.91 | 0.85 | 0.87 | 4,759 | 0.81 | 0.74 | 0.76 | 338 |
| Id | 1.00 | 0.98 | 0.99 | 341 | 1.00 | 1.00 | 1.00 | 15 |
| Methodology | 0.96 | 0.89 | 0.92 | 3,851 | 0.91 | 0.77 | 0.82 | 1,627 |
| Parameter | 0.83 | 0.76 | 0.79 | 3,003 | 0.78 | 0.68 | 0.73 | 1,345 |
| Result | 0.96 | 0.96 | 0.96 | 5,164 | 0.93 | 0.91 | 0.92 | 1,976 |
| Therapy | 0.97 | 0.85 | 0.90 | 1,273 | 0.33 | 0.50 | 0.40 | 2 |
| Weighted Mean | 0.93 | 0.88 | 0.90 | 19,055 | 0.88 | 0.80 | 0.84 | 5,605 |

### High recall on PICOS classification task shows utility of SURUS

To assess the utility of SURUS in the practice of systematic literature screening, we compared SURUS predictions to Cochrane-assigned PICOS labels for 8 randomly chosen interventional abstracts for the relevant therapeutic area. The results of the experiment are shown in Table 3. The overall F1 of SURUS during the utility assessment was 0.89. Most false positive predictions could be attributed to prediction of entities that made no appearance in the Cochrane “Characteristics of Studies” section. The high recall reflected a minimal risk of missing relevant elements of PICOS.

**Table 3.** Utility assessment metrics, matching SURUS predictions to mapped Cochrane extracts of elements of PICOS.

| PICOS label | TP | FP | FN | P | R | F1 |
| --- | --- | --- | --- | --- | --- | --- |
| Participants | 26 | 4 | 1 | 0.87 | 0.96 | 0.91 |
| Interventions / Comparisons | 32 | 3 | 2 | 0.91 | 0.94 | 0.93 |
| Outcomes | 27 | 9 | 1 | 0.75 | 0.96 | 0.84 |
| Study Design | 16 | 5 | 0 | 0.76 | 1 | 0.86 |
| Weighted Mean | 101 | 21 | 4 | 0.83 | 0.96 | 0.89 |

Abbreviations: TP: True Positive; FP: False Positive; FN: False Negative; P: Precision; R: Recall

## Discussion

In this paper, we evaluated a densely annotated and highly granular medical dataset for finetuning NLP text classification models. We compared the quality of multiple BERT model variants, fine-tuned on this dataset to identify named entities from clinical abstracts. Our measurements confirm that SURUS is capable of fine-grained classification and extraction of 25 different medically relevant categories, with a weighted mean F1 of 0.95 on interventional abstracts across 4 key therapeutic areas. The relatively high inter-annotator agreement (κ of 0.81) and the adequate out-of-domain performance of the fine-tuned underline the quality of the dataset. The high recall measured during the utility assessment demonstrate the value of SURUS to systematic literature reviewers in the screening process. The dataset and the annotation manual are available to the public and allow for expansion of the dataset for use in other domains.

To the best of our knowledge, of annotated medical NLP corpora published, the SURUS annotated dataset allows for the highest label prediction quality, for the largest diversity of clinical entity types. In addition, it shows the highest prediction quality of elements of PICOS as extracted by experts. This metric provides the key utility advantage of SURUS, granting high, time-saving opportunities to systematic literature reviewers with low risk of missing relevant elements of PICOS.

Current classification model alternatives typically focus on sentence or sentence clause classification, leaving much of the interpretation to the scientist performing the screening. In addition, mapping such text strands towards an ontology is laborious and inefficient. The fine-grained extraction of 25 labels allow SURUS to provide the reviewer with more detailed information on the PICOS element of studies in their selection. Important study features, such as information on drugs and treatments (0.95), elements of methodology (0.94) and disease (0.95) are predicted with high reliability, with limited variation between runs of the k-fold validation and in-domain therapeutic areas. Prediction quality in the current paper exceeds the current state-of-the-art prediction quality on other datasets focused on clinical studies such as EBM-PICO (0.73, PubmedBERT [22]), NICTA-PIBOSO (0.57-0.91, BioBERT [41]) and comparable to PubMedPICO (0.85-0.99, BioBERT [41]), recognizing more granular text spans and more label classes in the process. With the annotation manual and the test set (available on our git repository), the quality and feasibility of the SURUS annotation method can be reproduced. In addition, the SURUS is the only PICOS classification model on which the utility is assessed compared to mapped expert extractions, rather than annotation span comparison, which typically introduces a layer of subjectivity and inconsistency. When applying the trained system on abstracts in other therapeutic areas than the ones included, system performance remains sufficient. Even though the quality drops from 0.95 to 0.90, the most important classes for PICOS extraction retain high prediction quality. This signifies the utility of SURUS to systematic researchers specialized in any therapeutic area. Nevertheless, users are advised to produce a limited annotation set to include in the training procedure. This additional training dataset may be as small as 20 articles for out-of-domain therapeutic areas, considering the model quality already surpasses an F1 of 0.92 using 20% of the SURUS dataset as train dataset.

The prediction quality of SURUS falls off slightly for abstracts of observational studies compared to out-domain therapeutic area prediction (F1 of 0.84 vs 0.90). The discrepancy is likely because of the methodological and stylistic differences between study types. For example, in some observational studies diseases may be key study group differentiators, whereas in interventional studies, study groups are defined based on the therapeutic regimens received. In addition, there is large variety in writing style between different types of observational studies, which include study types such as diagnostic accuracy studies, cohort studies and case reports. Still, important NER class categories such as Disease and Methodology can relatively reliably be extracted from observational studies (F1 of 0.91 and 0.82, respectively). The current prediction quality offers perspective for additional fine-tuning efforts to improve the prediction quality of relevant medical labels in observational studies.

## Conclusion

Our findings show that the SURUS system is well-suited to classify 25 different medically relevant entity labels in interventional study abstracts with high prediction quality. Combined, its predictions can be used to extract elements of PICOS from clinical abstracts with high accuracy. Prediction quality is highest for articles on indications the system is trained on but remains considerable when applying SURUS to other indications. In addition, SURUS shows considerable practical utility when used to extract elements of PICOS from scientific abstracts, with very limited risk of failing to identify elements of PICOS.

## Data Availability

The dataset, annotation manual and model code are available upon request and will be published upon journal publication.

## Contributions

Investigation and writing of this work was done by Casper Peeters. Dr. Marianne Pouwer, Dr. Bart Westerman, Dr. Maikel Boot and Prof. Suzan Verberne co-authored the manuscript text. The SURUS annotation interface and the underlying BERT models were designed and maintained by Koen Vijverberg and Casper Peeters. The annotation manual was assembled by Casper Peeters.

## Acknowledgements

Khadija Ahmiane and Finn Bohte contributed to the assembly of the inter-annotator agreement set, Chayenne van Dongen assisted in the assembly of out-of-domain datasets. Finally, we would like to acknowledge all the master students who participated in the annotation process of the NER dataset.

## Funding

This work was funded by Medstone Holding BV, Almere, the Netherlands. Bart Westerman is supported by Health∼Holland under the grant called the toxicity atlas.

## Competing Interests

Casper Peeters and Koen Vijverberg are current employees of Medstone Holding BV, as was Dr. Marianne Pouwer at the time the study was conducted.

## Appendix

**Appendix table A.**
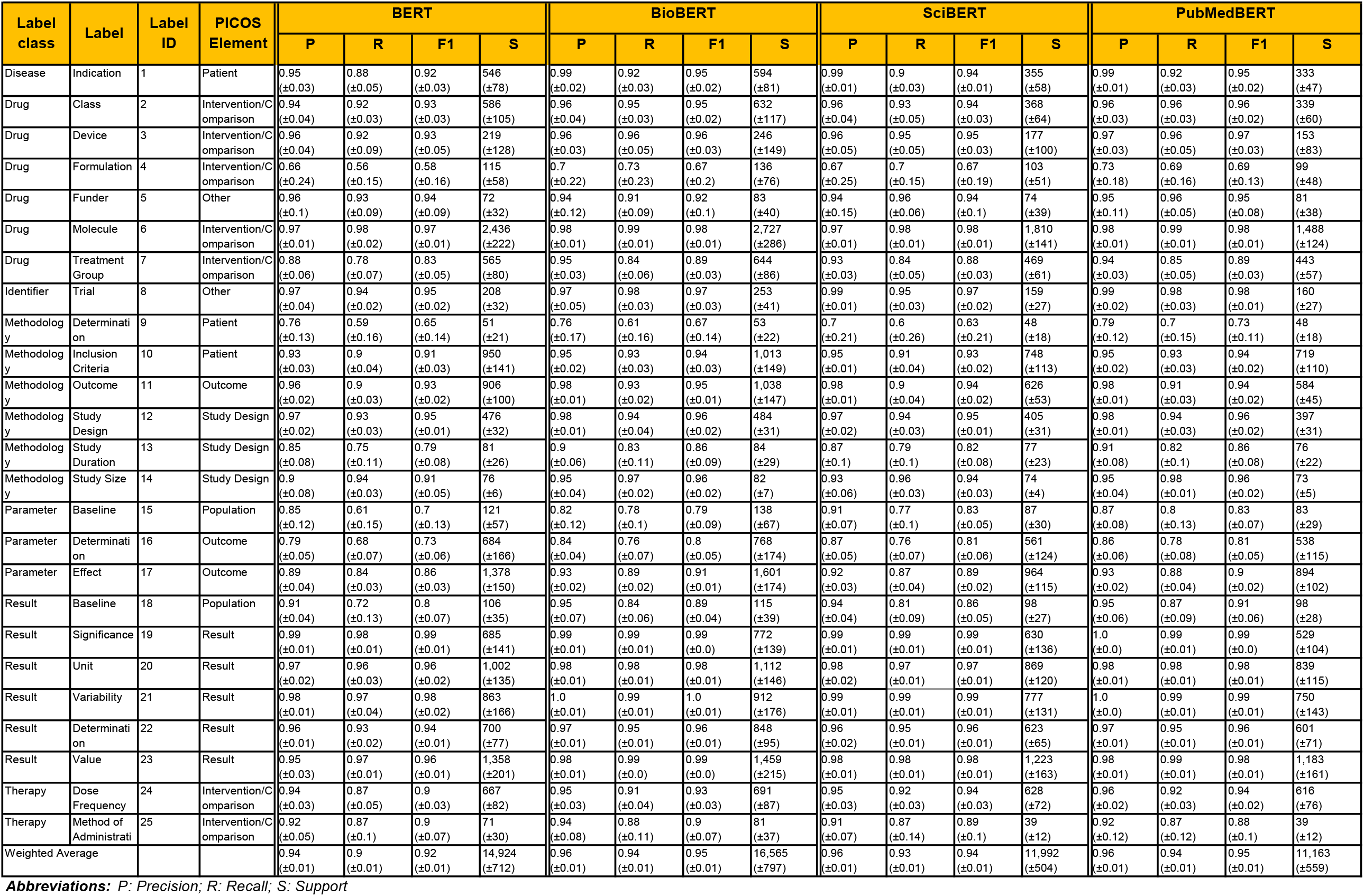
F1, precision, recall and support data for all annotation classes individually and the weighed average for all evaluation experiments.

| Label class | Label | Label ID | PICOS Element | BERT |  |  |  | BioBERT |  |  |  | SciBERT |  |  |  | PubMedBERT |  |  |  |
| --- | --- | --- | --- | --- | --- | --- | --- | --- | --- | --- | --- | --- | --- | --- | --- | --- | --- | --- | --- |
|  |  |  |  | P | R | F1 | S | P | R | F1 | S | P | R | F1 | S | P | R | F1 | S |
| Disease | Indication | 1 | Patient | 0.95<br>(±0.03) | 0.88<br>(±0.05) | 0.92<br>(±0.03) | 546<br>(±78) | 0.99<br>(±0.02) | 0.92<br>(±0.03) | 0.95<br>(±0.02) | 594<br>(±81) | 0.99<br>(±0.01) | 0.9<br>(±0.03) | 0.94<br>(±0.01) | 355<br>(±58) | 0.99<br>(±0.01) | 0.92<br>(±0.03) | 0.95<br>(±0.02) | 333<br>(±47) |
| Drug | Class | 2 | Intervention/Comparison | 0.94<br>(±0.04) | 0.92<br>(±0.03) | 0.93<br>(±0.03) | 586<br>(±105) | 0.96<br>(±0.04) | 0.95<br>(±0.03) | 0.95<br>(±0.02) | 632<br>(±117) | 0.96<br>(±0.04) | 0.93<br>(±0.05) | 0.94<br>(±0.03) | 368<br>(±64) | 0.96<br>(±0.03) | 0.96<br>(±0.03) | 0.96<br>(±0.02) | 339<br>(±60) |
| Drug | Device | 3 | Intervention/Comparison | 0.96<br>(±0.04) | 0.92<br>(±0.09) | 0.93<br>(±0.05) | 219<br>(±128) | 0.96<br>(±0.03) | 0.96<br>(±0.05) | 0.96<br>(±0.03) | 246<br>(±149) | 0.96<br>(±0.05) | 0.95<br>(±0.05) | 0.95<br>(±0.03) | 177<br>(±100) | 0.97<br>(±0.03) | 0.96<br>(±0.05) | 0.97<br>(±0.03) | 153<br>(±83) |
| Drug | Formulation | 4 | Intervention/Comparison | 0.66<br>(±0.24) | 0.56<br>(±0.15) | 0.58<br>(±0.16) | 115<br>(±58) | 0.7<br>(±0.22) | 0.73<br>(±0.23) | 0.67<br>(±0.2) | 136<br>(±76) | 0.67<br>(±0.25) | 0.7<br>(±0.15) | 0.67<br>(±0.19) | 103<br>(±51) | 0.73<br>(±0.18) | 0.69<br>(±0.16) | 0.69<br>(±0.13) | 99<br>(±48) |
| Drug | Funder | 5 | Other | 0.96<br>(±0.1) | 0.93<br>(±0.09) | 0.94<br>(±0.09) | 72<br>(±32) | 0.94<br>(±0.12) | 0.91<br>(±0.09) | 0.92<br>(±0.1) | 83<br>(±40) | 0.94<br>(±0.15) | 0.96<br>(±0.06) | 0.94<br>(±0.1) | 74<br>(±39) | 0.95<br>(±0.11) | 0.96<br>(±0.05) | 0.95<br>(±0.08) | 81<br>(±38) |
| Drug | Molecule | 6 | Intervention/Comparison | 0.97<br>(±0.01) | 0.98<br>(±0.02) | 0.97<br>(±0.01) | 2,436<br>(±222) | 0.98<br>(±0.01) | 0.99<br>(±0.01) | 0.98<br>(±0.01) | 2,727<br>(±286) | 0.97<br>(±0.01) | 0.98<br>(±0.01) | 0.98<br>(±0.01) | 1,810<br>(±141) | 0.98<br>(±0.01) | 0.99<br>(±0.01) | 0.98<br>(±0.01) | 1,488<br>(±124) |
| Drug | Treatment Group | 7 | Intervention/Comparison | 0.88<br>(±0.06) | 0.78<br>(±0.07) | 0.83<br>(±0.05) | 565<br>(±80) | 0.95<br>(±0.03) | 0.84<br>(±0.06) | 0.89<br>(±0.03) | 644<br>(±86) | 0.93<br>(±0.03) | 0.84<br>(±0.05) | 0.88<br>(±0.03) | 469<br>(±61) | 0.94<br>(±0.03) | 0.85<br>(±0.05) | 0.89<br>(±0.03) | 443<br>(±57) |
| Identifier | Trial | 8 | Other | 0.97<br>(±0.04) | 0.94<br>(±0.02) | 0.95<br>(±0.02) | 208<br>(±32) | 0.97<br>(±0.05) | 0.98<br>(±0.03) | 0.97<br>(±0.03) | 253<br>(±41) | 0.99<br>(±0.01) | 0.95<br>(±0.03) | 0.97<br>(±0.02) | 159<br>(±27) | 0.99<br>(±0.02) | 0.98<br>(±0.03) | 0.98<br>(±0.01) | 160<br>(±27) |
| Methodology | Determination | 9 | Patient | 0.76<br>(±0.13) | 0.59<br>(±0.16) | 0.65<br>(±0.14) | 51<br>(±21) | 0.76<br>(±0.17) | 0.61<br>(±0.16) | 0.67<br>(±0.14) | 53<br>(±22) | 0.7<br>(±0.21) | 0.6<br>(±0.26) | 0.63<br>(±0.21) | 48<br>(±18) | 0.79<br>(±0.12) | 0.7<br>(±0.15) | 0.73<br>(±0.11) | 48<br>(±18) |
| Methodology | Inclusion Criteria | 10 | Patient | 0.93<br>(±0.03) | 0.9<br>(±0.04) | 0.91<br>(±0.03) | 950<br>(±141) | 0.95<br>(±0.02) | 0.93<br>(±0.03) | 0.94<br>(±0.03) | 1,013<br>(±149) | 0.95<br>(±0.01) | 0.91<br>(±0.04) | 0.93<br>(±0.02) | 748<br>(±113) | 0.95<br>(±0.02) | 0.93<br>(±0.03) | 0.94<br>(±0.02) | 719<br>(±110) |
| Methodology | Outcome | 11 | Outcome | 0.96<br>(±0.02) | 0.9<br>(±0.03) | 0.93<br>(±0.02) | 906<br>(±100) | 0.98<br>(±0.01) | 0.93<br>(±0.02) | 0.95<br>(±0.01) | 1,038<br>(±147) | 0.98<br>(±0.01) | 0.9<br>(±0.04) | 0.94<br>(±0.02) | 626<br>(±53) | 0.98<br>(±0.01) | 0.91<br>(±0.03) | 0.94<br>(±0.02) | 584<br>(±45) |
| Methodology | Study Design | 12 | Study Design | 0.97<br>(±0.02) | 0.93<br>(±0.03) | 0.95<br>(±0.01) | 476<br>(±32) | 0.98<br>(±0.01) | 0.94<br>(±0.04) | 0.96<br>(±0.02) | 484<br>(±31) | 0.97<br>(±0.02) | 0.94<br>(±0.03) | 0.95<br>(±0.01) | 405<br>(±31) | 0.98<br>(±0.01) | 0.94<br>(±0.03) | 0.96<br>(±0.02) | 397<br>(±31) |
| Methodology | Study Duration | 13 | Study Design | 0.85<br>(±0.08) | 0.75<br>(±0.11) | 0.79<br>(±0.08) | 81<br>(±26) | 0.9<br>(±0.06) | 0.83<br>(±0.11) | 0.86<br>(±0.09) | 84<br>(±29) | 0.87<br>(±0.1) | 0.79<br>(±0.1) | 0.82<br>(±0.08) | 77<br>(±23) | 0.91<br>(±0.08) | 0.82<br>(±0.1) | 0.86<br>(±0.08) | 76<br>(±22) |
| Methodology | Study Size | 14 | Study Design | 0.9<br>(±0.08) | 0.94<br>(±0.03) | 0.91<br>(±0.05) | 76<br>(±6) | 0.95<br>(±0.04) | 0.97<br>(±0.02) | 0.96<br>(±0.02) | 82<br>(±7) | 0.93<br>(±0.06) | 0.96<br>(±0.03) | 0.94<br>(±0.03) | 74<br>(±4) | 0.95<br>(±0.04) | 0.98<br>(±0.01) | 0.96<br>(±0.02) | 73<br>(±5) |
| Parameter | Baseline | 15 | Population | 0.85<br>(±0.12) | 0.61<br>(±0.15) | 0.7<br>(±0.13) | 121<br>(±57) | 0.82<br>(±0.12) | 0.78<br>(±0.1) | 0.79<br>(±0.09) | 138<br>(±67) | 0.91<br>(±0.07) | 0.77<br>(±0.1) | 0.83<br>(±0.05) | 87<br>(±30) | 0.87<br>(±0.08) | 0.8<br>(±0.13) | 0.83<br>(±0.07) | 83<br>(±29) |
| Parameter | Determination | 16 | Outcome | 0.79<br>(±0.05) | 0.68<br>(±0.07) | 0.73<br>(±0.06) | 684<br>(±166) | 0.84<br>(±0.04) | 0.76<br>(±0.07) | 0.8<br>(±0.05) | 768<br>(±174) | 0.87<br>(±0.05) | 0.76<br>(±0.07) | 0.81<br>(±0.06) | 561<br>(±124) | 0.86<br>(±0.06) | 0.78<br>(±0.08) | 0.81<br>(±0.05) | 538<br>(±115) |
| Parameter | Effect | 17 | Outcome | 0.89<br>(±0.04) | 0.84<br>(±0.03) | 0.86<br>(±0.03) | 1,378<br>(±150) | 0.93<br>(±0.02) | 0.89<br>(±0.02) | 0.91<br>(±0.01) | 1,601<br>(±174) | 0.92<br>(±0.03) | 0.87<br>(±0.04) | 0.89<br>(±0.02) | 964<br>(±115) | 0.93<br>(±0.02) | 0.88<br>(±0.04) | 0.9<br>(±0.02) | 894<br>(±102) |
| Result | Baseline | 18 | Population | 0.91<br>(±0.04) | 0.72<br>(±0.13) | 0.8<br>(±0.07) | 106<br>(±35) | 0.95<br>(±0.07) | 0.84<br>(±0.06) | 0.89<br>(±0.04) | 115<br>(±39) | 0.94<br>(±0.04) | 0.81<br>(±0.09) | 0.86<br>(±0.05) | 98<br>(±27) | 0.95<br>(±0.06) | 0.87<br>(±0.09) | 0.91<br>(±0.06) | 98<br>(±28) |
| Result | Significance | 19 | Result | 0.99<br>(±0.01) | 0.98<br>(±0.01) | 0.99<br>(±0.01) | 685<br>(±141) | 0.99<br>(±0.01) | 0.99<br>(±0.01) | 0.99<br>(±0.0) | 772<br>(±139) | 0.99<br>(±0.01) | 0.99<br>(±0.01) | 0.99<br>(±0.01) | 630<br>(±136) | 1.0<br>(±0.0) | 0.99<br>(±0.01) | 0.99<br>(±0.0) | 529<br>(±104) |
| Result | Unit | 20 | Result | 0.97<br>(±0.02) | 0.96<br>(±0.03) | 0.96<br>(±0.02) | 1,002<br>(±135) | 0.98<br>(±0.01) | 0.98<br>(±0.01) | 0.98<br>(±0.01) | 1,112<br>(±146) | 0.98<br>(±0.02) | 0.97<br>(±0.01) | 0.97<br>(±0.01) | 869<br>(±120) | 0.98<br>(±0.01) | 0.98<br>(±0.01) | 0.98<br>(±0.01) | 839<br>(±115) |
| Result | Variability | 21 | Result | 0.98<br>(±0.01) | 0.97<br>(±0.04) | 0.98<br>(±0.02) | 863<br>(±166) | 1.0<br>(±0.01) | 0.99<br>(±0.01) | 1.0<br>(±0.01) | 912<br>(±176) | 0.99<br>(±0.01) | 0.99<br>(±0.01) | 0.99<br>(±0.01) | 777<br>(±131) | 1.0<br>(±0.0) | 0.99<br>(±0.01) | 0.99<br>(±0.01) | 750<br>(±143) |
| Result | Determination | 22 | Result | 0.96<br>(±0.01) | 0.93<br>(±0.02) | 0.94<br>(±0.01) | 700<br>(±77) | 0.97<br>(±0.01) | 0.95<br>(±0.01) | 0.96<br>(±0.01) | 848<br>(±95) | 0.96<br>(±0.02) | 0.95<br>(±0.01) | 0.96<br>(±0.01) | 623<br>(±65) | 0.97<br>(±0.01) | 0.95<br>(±0.01) | 0.96<br>(±0.01) | 601<br>(±71) |
| Result | Value | 23 | Result | 0.95<br>(±0.03) | 0.97<br>(±0.01) | 0.96<br>(±0.01) | 1,358<br>(±201) | 0.98<br>(±0.01) | 0.99<br>(±0.0) | 0.99<br>(±0.0) | 1,459<br>(±215) | 0.98<br>(±0.01) | 0.98<br>(±0.01) | 0.98<br>(±0.01) | 1,223<br>(±163) | 0.98<br>(±0.01) | 0.99<br>(±0.01) | 0.98<br>(±0.01) | 1,183<br>(±161) |
| Therapy | Dose Frequency | 24 | Intervention/Comparison | 0.94<br>(±0.03) | 0.87<br>(±0.05) | 0.9<br>(±0.03) | 667<br>(±82) | 0.95<br>(±0.03) | 0.91<br>(±0.04) | 0.93<br>(±0.03) | 691<br>(±87) | 0.95<br>(±0.03) | 0.92<br>(±0.03) | 0.94<br>(±0.03) | 628<br>(±72) | 0.96<br>(±0.02) | 0.92<br>(±0.03) | 0.94<br>(±0.02) | 616<br>(±76) |
| Therapy | Method of Administration | 25 | Intervention/Comparison | 0.92<br>(±0.05) | 0.87<br>(±0.1) | 0.9<br>(±0.07) | 71<br>(±30) | 0.94<br>(±0.08) | 0.88<br>(±0.11) | 0.9<br>(±0.07) | 81<br>(±37) | 0.91<br>(±0.07) | 0.87<br>(±0.14) | 0.89<br>(±0.1) | 39<br>(±12) | 0.92<br>(±0.12) | 0.87<br>(±0.12) | 0.88<br>(±0.1) | 39<br>(±12) |
| Weighted Average |  |  |  | 0.94<br>(±0.01) | 0.9<br>(±0.01) | 0.92<br>(±0.01) | 14,924<br>(±712) | 0.96<br>(±0.01) | 0.94<br>(±0.01) | 0.95<br>(±0.01) | 16,565<br>(±797) | 0.96<br>(±0.01) | 0.93<br>(±0.01) | 0.94<br>(±0.01) | 11,992<br>(±504) | 0.96<br>(±0.01) | 0.94<br>(±0.01) | 0.95<br>(±0.01) | 11,163<br>(±559) |

**Abbreviations:** P: Precision; R: Recall; S: Support

**Appendix table B.** Overview of WHO ICD-11 therapeutic areas included in the in-domain and out-domain datasets

| Annotation class category | Class description | Contribution to PICOS extraction |
| --- | --- | --- |
| **Disease** | Any reference to disease, independent of relevance to described study | Patient |
| **Drug** | Any reference to treatments | Intervention/Comparison |
| **Identifier** | Trial identifiers of clinical trials | - |
| **Methodology** | Any reference to elements of design, inclusion criteria, outcomes measured, size or duration of the described study | Patient, Outcome, Study design |
| **Parameter** | References to outcomes of the study of which results are described | Outcome |
| **Result** | Any reference to a result, result description or significance | - |
| **Therapy** | Any reference to treatment dosing and administration frequency | Intervention/Comparison |

**Appendix table C.**
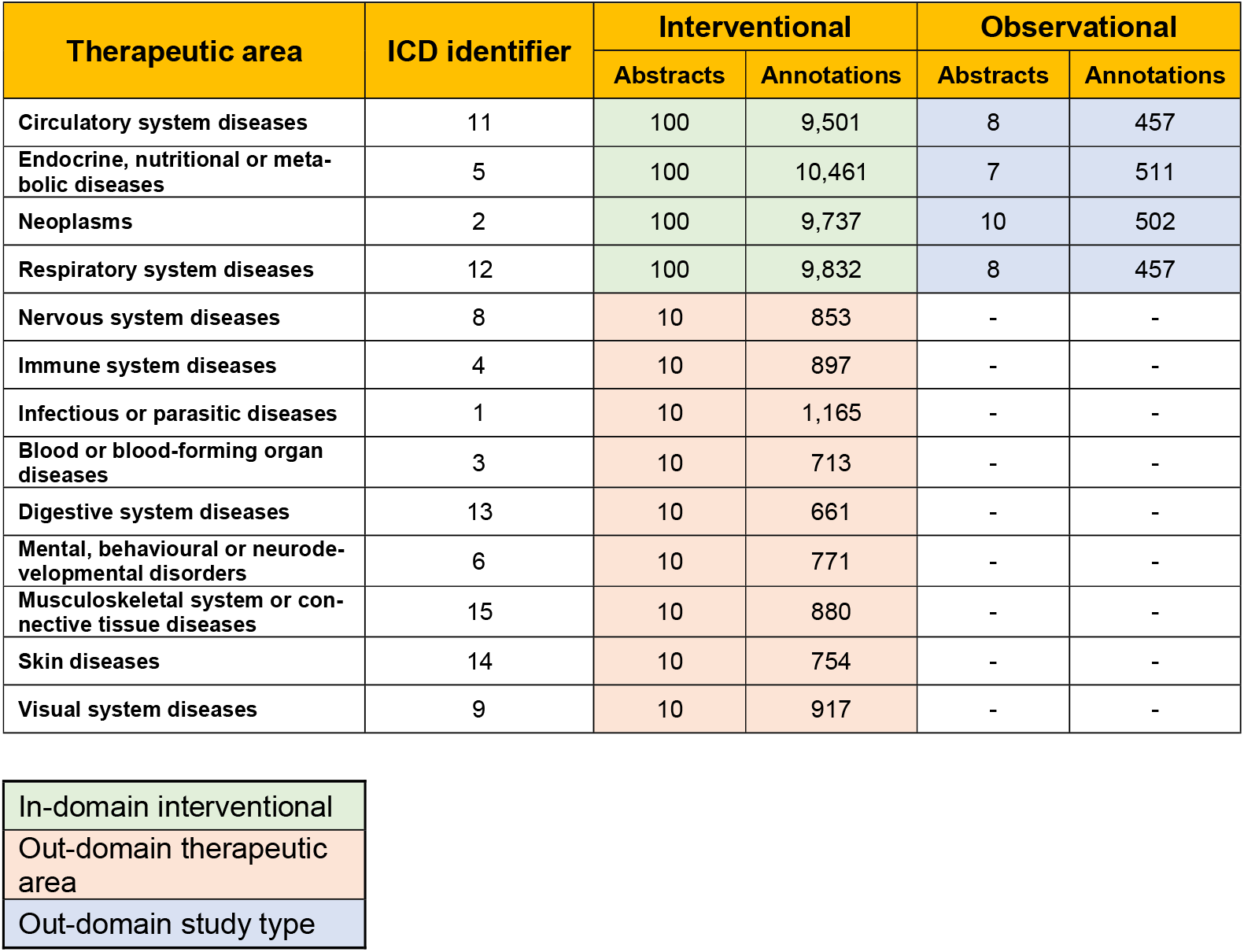
Number of articles and annotations included in every model assessment described.

| Therapeutic area | ICD identifier | Interventional |  | Observational |  |
| --- | --- | --- | --- | --- | --- |
|  |  | Abstracts | Annotations | Abstracts | Annotations |
| Circulatory system diseases | 11 | 100 | 9,501 | 8 | 457 |
| Endocrine, nutritional or metabolic diseases | 5 | 100 | 10,461 | 7 | 511 |
| Neoplasms | 2 | 100 | 9,737 | 10 | 502 |
| Respiratory system diseases | 12 | 100 | 9,832 | 8 | 457 |
| Nervous system diseases | 8 | 10 | 853 | - | - |
| Immune system diseases | 4 | 10 | 897 | - | - |
| Infectious or parasitic diseases | 1 | 10 | 1,165 | - | - |
| Blood or blood-forming organ diseases | 3 | 10 | 713 | - | - |
| Digestive system diseases | 13 | 10 | 661 | - | - |
| Mental, behavioural or neurodevelopmental disorders | 6 | 10 | 771 | - | - |
| Musculoskeletal system or connective tissue diseases | 15 | 10 | 880 | - | - |
| Skin diseases | 14 | 10 | 754 | - | - |
| Visual system diseases | 9 | 10 | 917 | - | - |

|  |
| --- |
| In-domain interventional |
| Out-domain therapeutic area |
| Out-domain study type |

## Footnotes

1 https://www.cochranelibrary.com/about/pico-search

2 https://github.com/jind11/NICTA-PIBOSO-Dataset

3 https://paperswithcode.com/dataset/pubmed-pico-element-detection-dataset

4 https://ebm-nlp.herokuapp.com/

5 https://pubmed.ncbi.nlm.nih.gov/

6 https://icd.who.int/browse11/l-m/en

7 https://huggingface.co/transformers/model_doc/bert.html#bertto-kenizer

8 https://huggingface.co/bert-base-uncased

## Notes

### Summary of Updates

Minor correction of abstract sentence structure; Correction of table order.

